# Prevalence of class A ESBL, class B and D carbapenemase encoding genes (CTX-M, TEM, SHV, NDM, IMP, OXA-48) in gram-negative bacterial pathogens isolated from various clinical samples collected from northern region of United Arab Emirates

**DOI:** 10.1101/2024.01.26.24301841

**Authors:** Premalatha Ragupathi, Vaneezeh Khamisani, Aisha Fadila Sadiq, Mariam Aylu Mobiddo, Nazeerullah Rahamathullah, Sovan Bagchi, Ala Kamian, Nasir Parwaiz, Mohammad Shahid Akhtar

## Abstract

**Objective:** The aim of this study was to assess the prevalence of class A ESBL, class B and D carbapenemase encoding genes (blaCTX-M, blaTEM, blaSHV, blaNDM, blaIMP and blaOXA-48) in gram-negative bacterial pathogens isolated from various clinical samples collected from northern region of UAE.

**Methods:** A laboratory based experimental study was conducted from October 14, 2021, to June 30, 2022. A total of 3670 various clinical samples (urine, blood, pus, and sputum) were obtained from patients attending the in and out-patient departments of Thumbay University Hospital and various other hospitals of Northern Emirates of UAE, processed for routine bacterial culture examination and determined their antibiotic sensitivity pattern especially ESBL and carbapenem resistance using MicroScan Neg Breakpoint Combo 50 panel. Molecular detection of class A ESBL, class B and D carbapenemase encoding genes (blacTX-M, blaTEM, blaSHV blaNDM, blaIMP & blaOXA-48) were performed on the ESBL and carbapenemase producing gram negative bacteria. Partial genomic sequencing and phylogenetic tree analysis was performed on the most predominant ESBL gene blaCTX-M.

**Results:** In total, 1098 bacterial cultures were obtained, of which 833 were gram negative bacteria and among them, 209 (25.1%) were ESBL producers and 40 (4.8%) were both ESBL and carbapenemase producers. In all the 249 bacterial isolates, the ESBL and carbapenemase encoding genes (*bla_CTX-M,_ bla_TEM_*, *bla_SHV_*, *bla_NDM_* & *bla_IMP_*) were detected except *bla_OXA-48_*. The *bla_CTX-M_ (n=72)* was the predominantly harbored gene followed by *bla_TEM_ (n=56)* and *bla_SHV_ (n=29),* mostly detected in *E. coli (n=157)*, then *K. pneumoniae (n=41), P. aeruginosa (n=18)* & *A. baumannii (n=7).* The *bla_NDM_* & *bla_IMP_* were detected in *K. pneumoniae (n=4), A. baumannii (n=3)* and *P. aeruginosa (n=2)* for *bla_IMP_*. The gene combinations such as *bla_CTX-M+TEM_ (n=15)*, *bla_CTX-M+SHV_ (n=15)*, *bla_CTX-M+TEM_*+*bla_SHV_ (n=6)* were co-harbored in 36 *E. coli* and in the *K. pneumoniae*, the *bla_CTX-M+TEM_ (n=15)*, *bla_CTX-M+SHV_ (n=15)*, *bla_TEM_*_+*SHV*_ *(n=3), bla_TEM+NDM_ (n=1)* were detected. The *bla_CTX-M+TEM,_ bla_CTX_-_M+SHV_* & *bla_CTX-M+TEM+SHV+IMP_* were detected in one *P. aeruginosa* and in one *A. baumannii,* the *bla_TEM+NDM_, bla_TEM+IMP_* & *bla_CTX-M+TEM+SHV+IMP_* were detected.

**Conclusion:** The highest bacterial culture positivity was detected in the urine samples. 25.1% of the Gram-ve bacteria were only ESBL producers and 4.8% were both the ESBL & carbapenemase producers. 44.5% of ESBL & carbapenemase producing bacteria harbored the *bla_CTX-M_* and mostly detected in the *E. coli,* then *K. pneumoniae* and *P. mirabilis.* Multiple gene combinations were detected in 5% *P. aeruginosa,* 10% *A. baumannii* and 3.82% *E. coli*. This finding highlights the importance of molecular detection of ESBL & carbapenemase producing genes to emphasize the monitor and control the development of multi drug resistant Gram-ve bacterial pathogens.

## Introduction

Antibiotic-resistant bacterial infections are now a serious problem worldwide and have become a global threat to public health, animal production, and environmental health. Globally, more than 7 million people die every year from multidrug resistant bacterial infection. It is estimated that 10 million people will die from antimicrobial resistant (AMR) infections by 2050.[1, 2] Over prescribing antibiotics, irresponsible usage and not adhering to prescription are some of the leading causes of development of antibiotic resistance.[3, 4] Gram-negative bacteria are more prone to develop antibiotic resistance than other bacteria and due to the enhanced use of beta-lactam antibiotics, the bacterial pathogens have developed to produce the enzyme beta lactamase using plasmid which counteract the beta lactam compound. [5, 6]

The rapid spread of antibiotic resistance genes among the gram-negative bacteria facilitated by the mobile genetic elements; plasmids and transposons are responsible for resistance towards a broad spectrum of antibiotics and have established the pivotal role in the dissemination of AMR.[7] Spreading of the plasmid mediated beta lactamase in gram negative bacterial pathogens is known as extended spectrum of beta lactamase (ESBL). Molecular classification of the β-lactamases divided into A, B, C, D enzymes [8, 9]. The class A enzyme producing genes are *bla_TEM_*, *bla_SHV_* and *bla_CTX-M_* [10]. The *bla_TEM_* is commonly found in Gram-negative bacteria [11, 12] and the gene *bla_CTX-M_* produces an enzyme called cefotaximase that inhibits the activity of the cefotaxime and as of March 2019, over 172 CTX-M types have been identified and described [13–15]. The gene *bla_SHV_*and *bla_TEM_* are structurally similar and as of February 2018, over 223 TEM and 193 SHV types have been listed in public databases [16].

The ESBL producers have emerged as a major source of resistance among the gram-negative bacteria, more predominantly in the *Escherichia coli* and *Klebsiella pneumoniae*; thereby becoming a global concern. [17] *K. pneumoniae* strains that produce ESBLs and carbapenemase have a worldwide distribution and cause multidrug-resistant (MDR) infections in many countries. [7, 18, 19]. Carbapenems are one of the few drugs that are useful for the treatment of MDR gram-negative bacterial infections. The emergence of carbapenem resistance in *Enterobacteriaceae* family is a serious public health concern due to the large spectrum of resistant genes and lack therapeutic options.[20,21] Carbapenem-hydrolyzing β-lactamases belonging to molecular class A (e.g., KPC, GES, IMI, SME), class B (e.g. IMP, VIM, NDM, GIM) and class D (e.g. OXA-23 & OXA-48) and are the main source of antibiotic resistance in *Enterobacteriaceae*.[20–22]

Furthermore, ESBL developing species show co-resistance to many other groups of antibiotics and limiting therapeutic options. As a result, in recent past, emergence of multidrug resistant (MDR) gram-negative bacterial infections have increased significantly and it is mainly acquired by ESBLs. [6,7, 23] Nevertheless there is still a scarcity of data on the prevalence of ESBLs and carbapenemase producing genes in patients’ samples in the UAE. Although some data have been published from this region and neighbouring countries, limited number of studies are available on the genes *bla_TEM_*, *bla_SHV_*, *bla_CTX-M_*, *bla_NDM_, bla_IMP_* and *bla*_OXA-48_ but no detailed studies on the prevalence and harboring of multiple genes in the MDR bacteria have yet been performed in the UAE. So, the current research focuses on the prevalence of class A ESBL (*bla_TEM_*, *bla_SHV_* & *bla_CTX-M_*) and class B & D carbapenemase encoding genes (*bla_NDM_, bla_IMP,_ bla*_OXA-48_) in gram-negative bacterial pathogens isolated from various clinical samples collected from northern region of UAE. Determining the prevalence of class A ESBL and class B & D carbapenemase producing genes in gram-negative bacterial pathogens is critical for effective treatment of infections caused by the MDR organisms especially in ICU and in-patients with co-morbidity cases.

## Methods

### Ethical Approval

The present study was approved by the Institutional Review Board (IRB) of Gulf Medical University, Ajman, UAE. The approval letter from the IRB, dated October 14, 2021, indicated that the study had already received approval in the IRB meeting of September 2021 (Ref.no.IRB/COM/STD/98/Oct.2021). The study design and protocol followed the Good Clinical Practice (GCP) guidelines 2021, National Drug Abuse Treatment Clinical Trials Network.

### Study Design

A laboratory based experimental study was conducted on various clinical samples such as urine, blood, pus, and sputum received from various hospitals of Northern Emirates of the UAE to the Department of Microbiology at Thumbay Laboratory, Thumbay University Hospital (TUH), Ajman, UAE.

### Data collection & Clinical sample processing

In accordance with the IRB approval, the patients’ clinical samples and other details were collected from Thumbay Laboratory, TUH, from September 02, 2021, to June 20, 2022. But laboratory analysis was performed following receipt of the IRB approval from October 14, 2021, to June 30, 2022.

### Study populations

All the clinical samples were received and collected from the patients who are attending internal medicine out-patient and in patient department of TUH and other hospitals of Northern Emirates of the UAE. All the samples were received by the phlebotomy department and sent to the Microbiology department at Thumbay Laboratory, TUH to culture bacterial pathogens, staining and antibiotic sensitivity test of the isolated pathogens, and molecular detection of class A ESBL genes *bla_CTX-M_, bla_SHV_, bla_TEM_*and carbapenemase producing genes *bla_NDM_, bla_IMP_,* and *bla_OXA-48_*. Upon arrival, the samples were processed for further laboratory tests.

### Bacterial isolation and staining

The clinical samples were plated on MacConkey’s agar to isolate the bacterial colony belongs to the *Enterobacteriaceae* family, Blood agar for gram positive bacterial species, Cysteine-Lactose Electrolyte Deficient (CLED) agar for urinary tract pathogens and Cetrimide Agar for *Pseudomonas* species isolation. All the culture plates were incubated overnight at 37°C for 18-24 hours. After the isolation of bacterial colonies from the different culture media, they were stained by Gram staining to identify them whether gram-negative or gram-positive bacteria.

### Bacterial identification and antibiotic sensitivity test

MicroScan WalkAway automated detection system was used to identify bacterial pathogens and to determine their ESBL and MDR nature. The bacterial inoculum was prepared from single colonies of isolated bacteria by using the Prompt™ Inoculation System-D (Beckman Coulter, Brea, CA, USA), consisting of a wand designed to hold a specific quantity of bacteria and a 30 ml Prompt™ Inoculation Water bottle (BECKMAN COULTER, Brea, CA, USA). The inoculum was suspended thoroughly, poured in to a RENOK inoculator tray and then transferred in to MicroScan Neg Breakpoint Combo 50 panel for Gram-negative organism (BECKMAN COULTER) using RENOK inoculator (BECKMAN COULTER). The plate was incubated at 16 – 18°C in MicroScan WalkAway automated system DxM 1096 (BECKMAN COULTER). Table 1 & 1a demonstrate various tests performed on the MicroScan Neg Breakpoint Combo 50 panel. The Beckman Coulter MicroScan LabPro software WalkAway 9631049 was used to evaluate the gram-negative bacterial pathogens on their biochemical utilization and antibiotic sensitivity pattern especially ESBL and carbapenem resistant strains. The details of the MicroScan Neg Breakpoint Combo 50 panel for Gram-negative organisms are given in <u>Supplementary table1a</u> & <u>1b</u>.

**Table 1.**
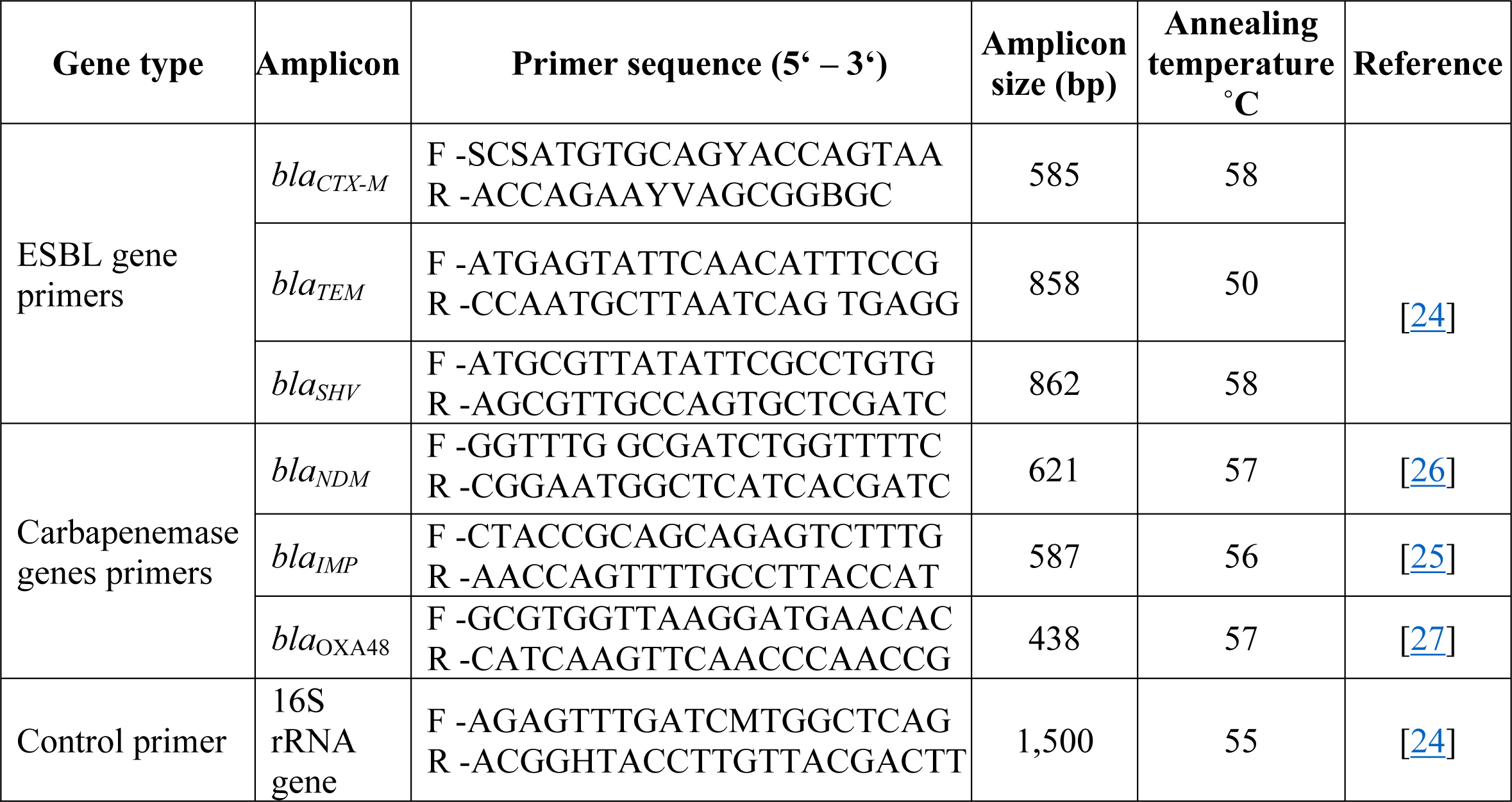
PCR primers and their specifications.

### Detection of Class A ESBL and Class B & D carbapenemase encoding genes

After the identification of ESBL and MDR strains, they were sub-cultured in sterile double strength Muller Hinton broth at 37°C for 24 hours for bacterial DNA extraction with the G-spin^TM^ for Genomic DNA extraction kit (iNtRON). The DNA extracts positive for the 16SrRNA gene in PCR assays were processed for the presence of the class A ESBL genes *bla_SHV_*, *bla_CTX-M_*, *bla_TEM_*and carbapenem resistant genes *bla_NDM_*, *bla_IMP,_* and *bla_OXA-48_* with consensus primers in a Veriti^TM^ 96-well Thermal cycler (Applied Bio-systems^TM^). The PCR assays were performed with Thermoscientific PCR Master mix kit (ThermoFisher). The PCR primers and their specifications are provided in the Table 1. Primers of all the class A ESBL and carbapenem resistant genes were synthesized by e-oligos, Gene Link^TM^, NY, USA. The PCR products underwent agarose gel electrophoresis to separate the fragments of DNA and visualized the target genes which are pointed in Fig 1.

**Fig 1.**
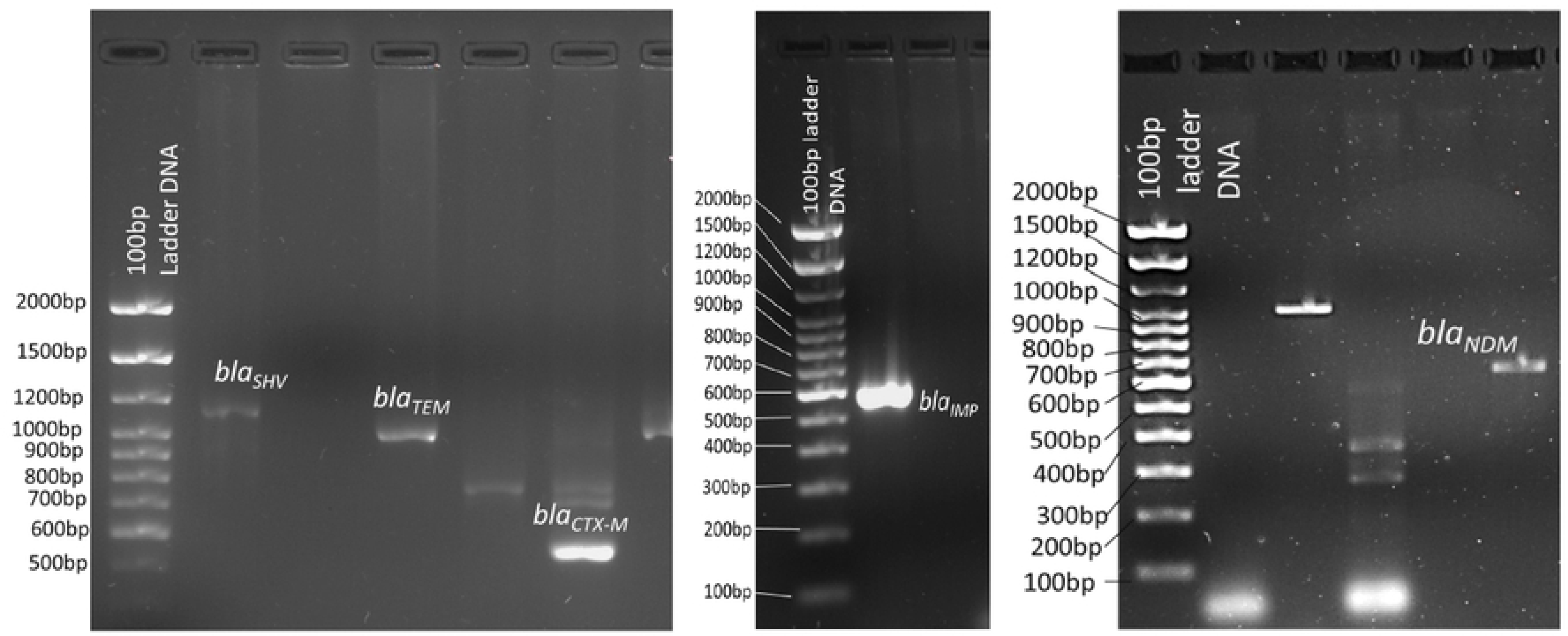
Detection of *bla_CTX-M_, bla_TEM_, bla_SHV_, bla_IMP_* & *bla_NDM_*bands in 1% agarose gel.

### Genomic sequencing and phylogenetic tree relationships of the *bla_CTX-M_*

Partial gene sequencing of the *bla_CTX-M_* of the *E. coli* was done by Sanger sequencing method and the sequenced product was compared with existing gene seqences in GenBank, NCBI. The cladogram of the phylogenetic tree relationship of the *bla_CTX-M_* was constructed using Mega 11 software. It was constructed using the Dice coefficient with a 1% tolerance limit and 1% optimization. Cluster relatedness of collected isolates with ≥ 70% similarity was considered to indicate an identical pattern type [28, 29].

### Statistical Analysis

Data were analyzed using the statistical package SPSS software version 28.0 (SPSS Inc., Chicago, IL, USA). Descriptive data were represented as mean ± SD. Pearson’s chi-square test was performed in tables 3 and 4 with *p*-value <0.05 considered as statistically significant.

**Table 2.** No. of various clinical samples for positive bacterial growth.

| Clinical samples<br>$n= 3670$<br>(%) | No. of samples for positive bacterial growth<br>$n=1098$ (29.92%) | | No. of samples for negative bacterial growth<br>$n=2572$<br>(70.08%) | Pearson's Chi-square test |
| --- | --- | --- | --- | --- |
| | Gram negative bacterial growth<br>$n= 833$<br>(75.86%) | Gram positive bacterial growth<br>$n=265$<br>(24.14%) | | |
| **Urine**<br>$n= 1970$<br>(53.7%) | 503<br>(60.4%) | 37<br>(13.96%) | 1430<br>(55.59%) | <0.0001 |
| **Blood**<br>$n= 879$<br>(23.93%) | 48<br>(5.8%) | 71<br>(26.8%) | 760<br>(29.54%) | |
| **Pus**<br>$n= 469$<br>(12.8%) | 111<br>(13.3%) | 142<br>(53.58%) | 216<br>(8.39%) | |
| **Sputum**<br>$n= 352$<br>(9.6%) | 171<br>(20.53%) | 15<br>(5.66%) | 166<br>(6.45%) | |

**Table 3.** Total no. of Gram-ve bacteria isolated from various clinical samples.

| Gram negative bacterial isolates | Total no. of bacteria (n=833) |  |  |  | Pearson's Chi-square test |
| --- | --- | --- | --- | --- | --- |
|  | Urine<br>n=503 (%) | Blood<br>n=48 (%) | Pus<br>n=111 (%) | Sputum<br>n=171(%) |  |
| *E. coli* | 354 (70.37%) | 8 (16.66%) | 25 (22.52%) | 7 (4.1%) | <0.0001 |
| *K. pneumoniae* | 116 (23.06%) | 15 (31.25%) | 35 (31.53%) | 34 (19.9%) |  |
| *P. aeruginosa* | 15 (2.98%) | 9 (18.75%) | 19 (17.11%) | 86 (50.29%) |  |
| *P. mirabilis* | 12 (2.39%) | 0 | 24 (21.62%) | 4 (2.34%) |  |
| *A. baumannii* | 6 (1.19%) | 16 (33.33%) | 8 (7.21%) | 40 (23.4%) |  |

**Table 4.**
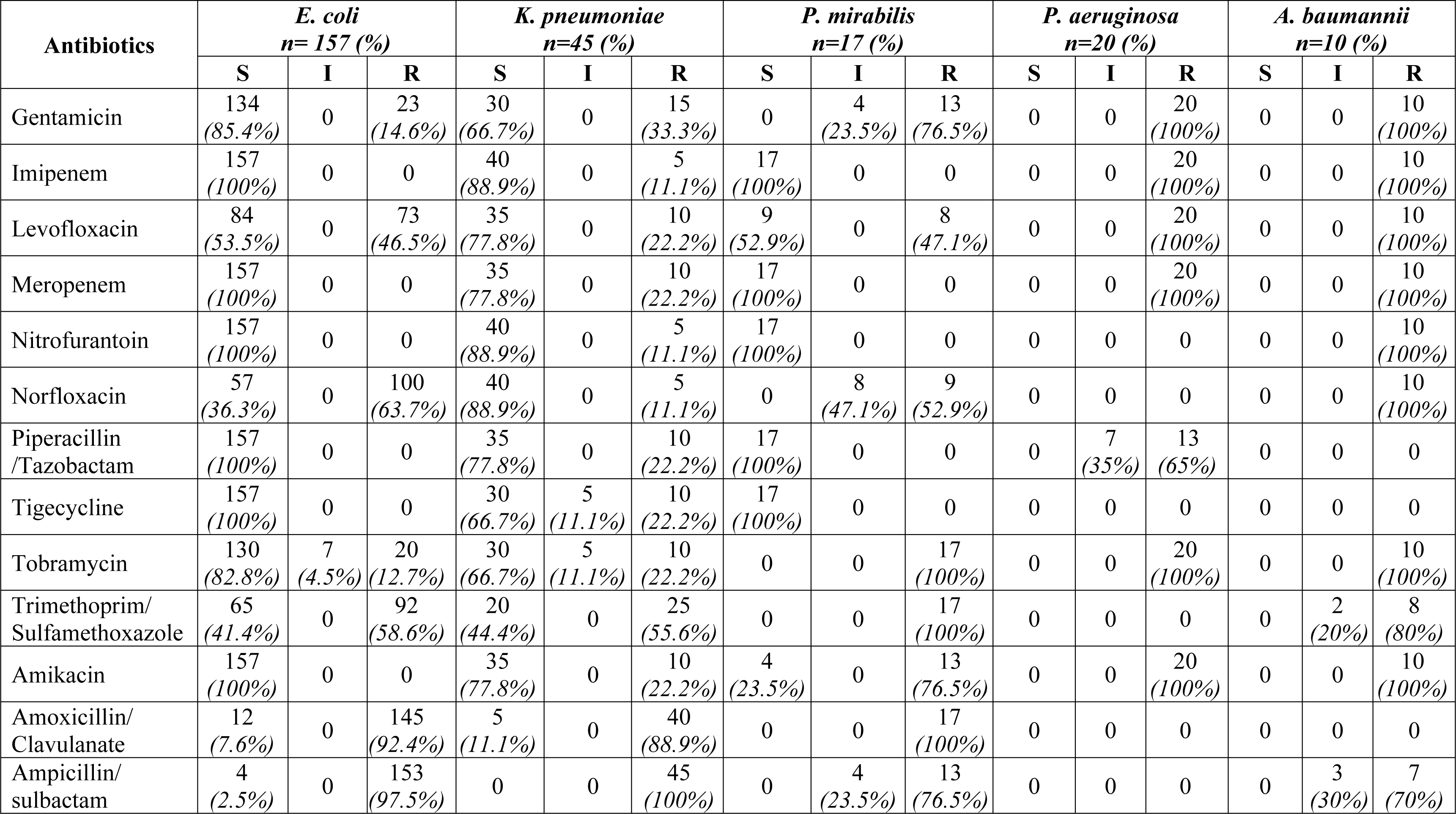
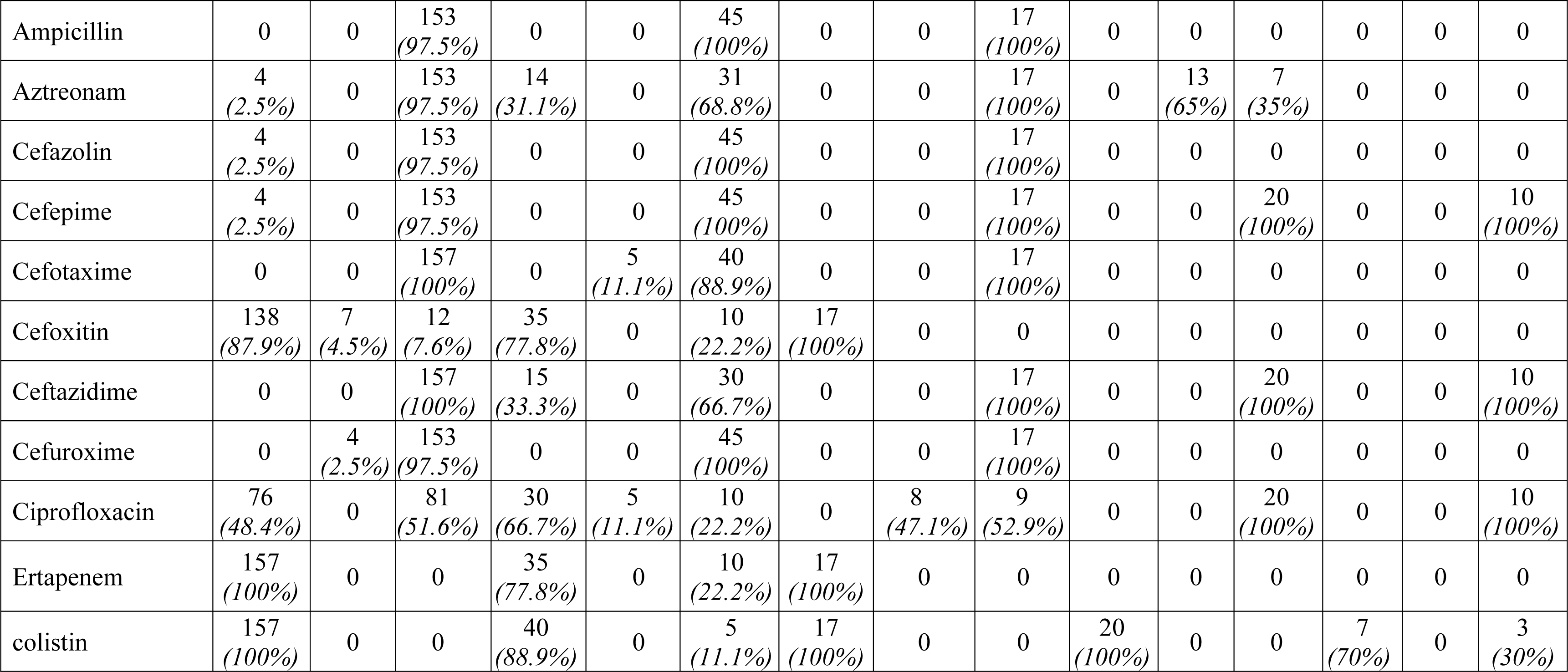
Antibiotic sensitivity pattern of *E. coli, K. pneumoniae, P. aeruginosa, P. mirabilis* and *A. baumannii* isolated from various clinical samples.

## RESULTS

### Bacterial isolation and staining

A total of 3670 samples were collected and processed for the routine bacterial culture. Among them, pus (n=469;12.7%), blood (n=879;24%), sputum (n=352;9.6%), urine (n=1970; 53.7%) samples were received, out of which, 1098 samples (29.9%) were positive and 2572 (70.1%) were negative for the bacterial culture. In that culture positive samples, the highest bacterial culture positivity was detected in the urine samples (49.1%; n=540), then in the pus (23%; n=253), blood (10.8%; n=119) and sputum (16.9%; n=186). In the 10986 culture positive samples, 833 (75.9%) were gram-negative and 265 (24.1%) were gram-positive bacteria and their details are given in Table 2; Fig 2.

**Fig 2.**
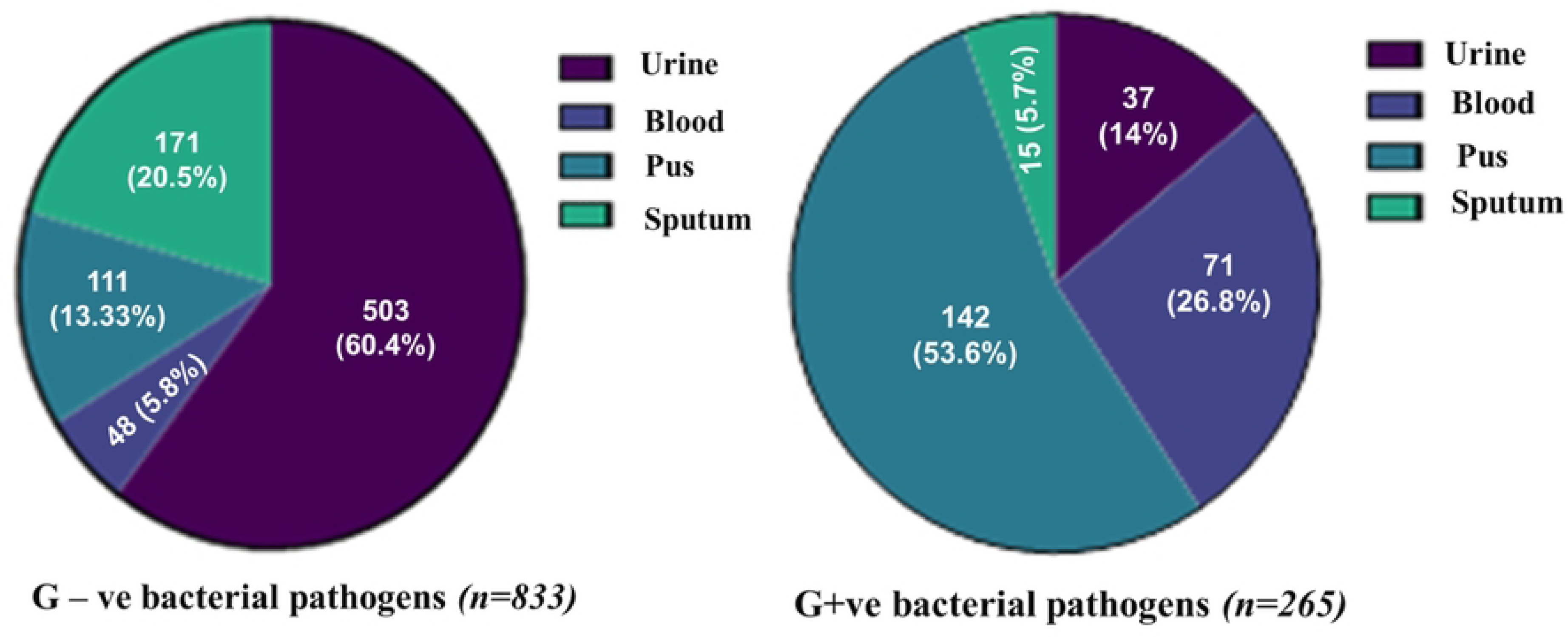
No. of Gram-ve and gram +ve bacteria isolated from various clinical samples.

### Bacterial identification and antibiotic susceptibility test

Among the gram-negative bacterial isolates, 249 (29.9%) were ESBL (*n=209*; 25.1%) and both ESBL & carbapenemase producers (*n=40*; 4.8%). Among the 249 bacterial isolates, 157 (63.1%) *E. coli*, 45 (18.1%) *K. pneumoniae*, 20 (8.0%) *Pseudomonas aeruginosa*, 17 (6.8%) *Proteus mirabilis,* and 10 (4.0%) *Acinetobacter baumanii* were encountered and provided in Table 3; Fig 3.

**Fig 3.**
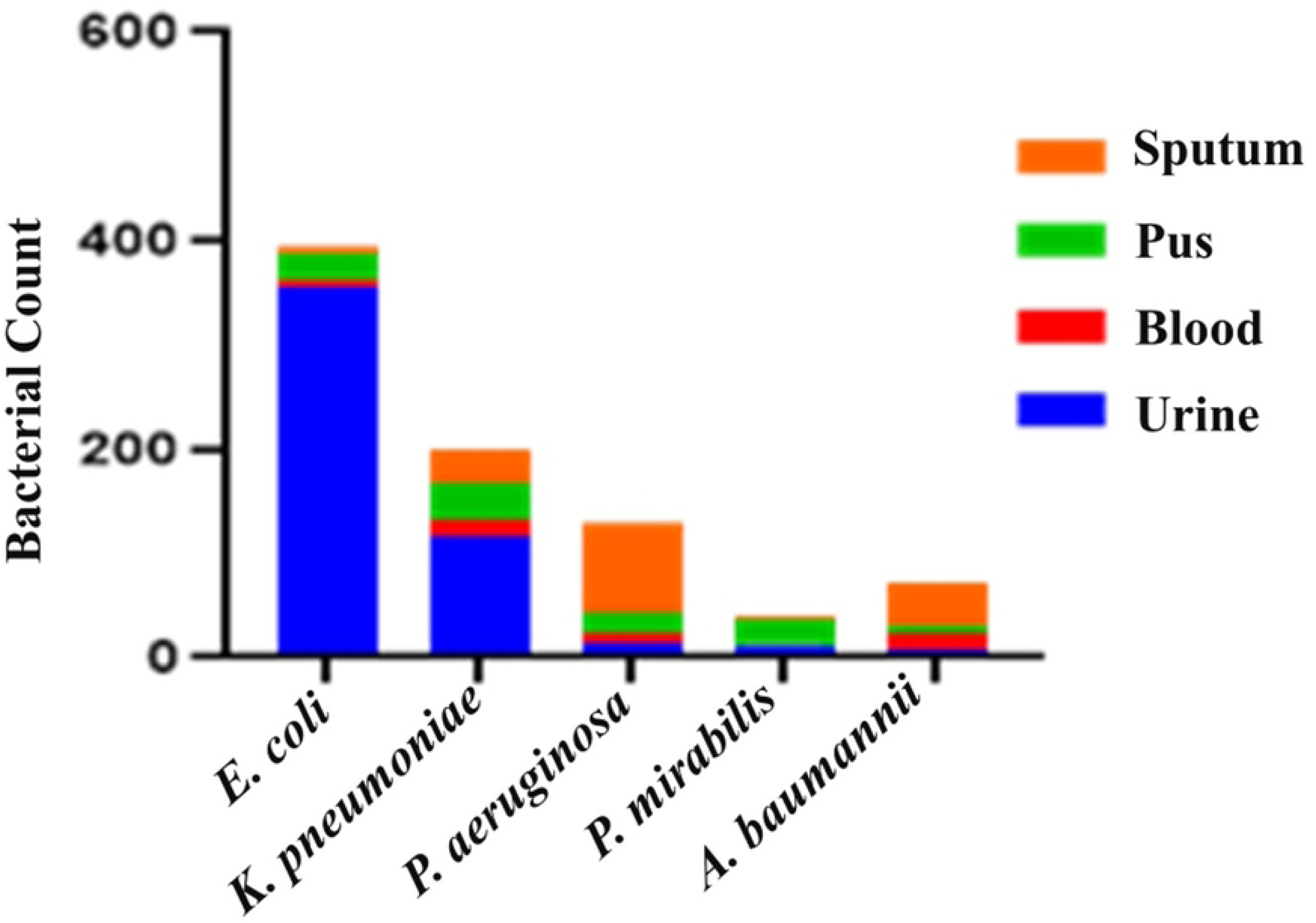
Total no. of *E. coli, K. pneumoniae, P. aeruginosa, P. mirabilis* and *A. baumannii* in various clinical samples.

The ESBL *E. coli* (*n*=157) had shown 100% resistance to cefotaxime and ceftazidime, high - medium resistance rates to Ampicillin/sulbactam, ampicillin, Aztreonam, cefazolin, cefepime, cefuroxime, (97.5%) & Amoxicillin+clavulanate (92.5%), norfloxacin (63.7%), trimethoprim/Sulfamethoxazole (58.6%), ciprofloxacin (51.6%) and levofloxacin (46.5%). Lower resistance rate to gentamycin (14.7%), tobramycin (12.7%) and least resistance rate to cefoxitin (7.6%) and absolute sensitivity to imipenem, meropenem, nitrofurantoin, piperacillin/tazobactam, tigecycline, amikacin, ertapenem, colistin. In the *K. pnuemoniae* isolates (*n=45*), all of them shown 100% resistant to the antibiotics: ampicillin, ampicillin + sulbactam, aztreonam, cefazolin, cefepime, and cefuroxime. 89% of them were resistant to amoxycillin + clavulanate and cefotaxime, whereas 66.7% - 55.6% of them were resistant to Ceftazidime, nitrofurantoin, norfloxacin, trimethoprim/sulfamethoxazole. 33.3% of the bacteria were resistant to the drug gentamycin. Nearly 22.2% of the bacteria were resistant to 9 drugs: levofloxacin, meropenem, pipracillin+tazobactum, tigecycline, tobramycin, Amikacin, cefoxitin, ertapenem and ciprofloxacin. 11.1% resistant to colistin, imipenem, norfloxacin. None of the *K. pneumoniae* had shown 100% sensitive to the above-mentioned drugs. In case of ESBL producing *P. mirabilis* (*n*=17), all were resistant to the following 10 antibiotics: tobramycin, amoxycillin+clavulanate, ampicillin, aztreonam, cefepime, cefazolin, cefotaxime, cefuroxime, trimethoprim/sulfamethoxazole and ceftazidime. 76.5% of them were shown resistance to the drugs gentamycin & amikacin. 52.9% were resistant to the drug norfloxacin, ciprofloxacin and levofloxacin. The bacteria had shown intermediate resistance to the following drugs norfloxacin, ciprofloxacin, gentamycin and ampicillin+sulbactam and they were 100% sensitive to imipenem, meropenem, nitrofurantoin, piperacillin/tazobactam, Tigecycline, cefoxitin, ertapenem & colistin. The bacteria *P. aeruginosa* (*n*=20) had shown 100% resistance to the drugs gentamycin, imipenem, levofloxacin, meropenem, tobramycin, amikacin, cefepime, ceftazidime and ciprofloxacin, 65% and 35% of them were resistant to the drugs pipracillin+tazobactum and Aztreonam respectively and none of them were 100% sensitive to any antibiotics which are used in the test except colistin. Similarly, the bacteria *A. baumannii* (n=10) had shown 100% resistance to 11 different drugs, those were amikacin, cefepime, ciprofloxacin, gentamycin, imipenem, levofloxacin, meropenem and ceftazidime. 80% of the strains were resistant to ampicillin + sulbactam, trimethoprim/sulfamethoxazole and tobramycin, and 30% of them were resistant to colistin. The details are given in Table 4; Fig 4.

**Fig 4.**
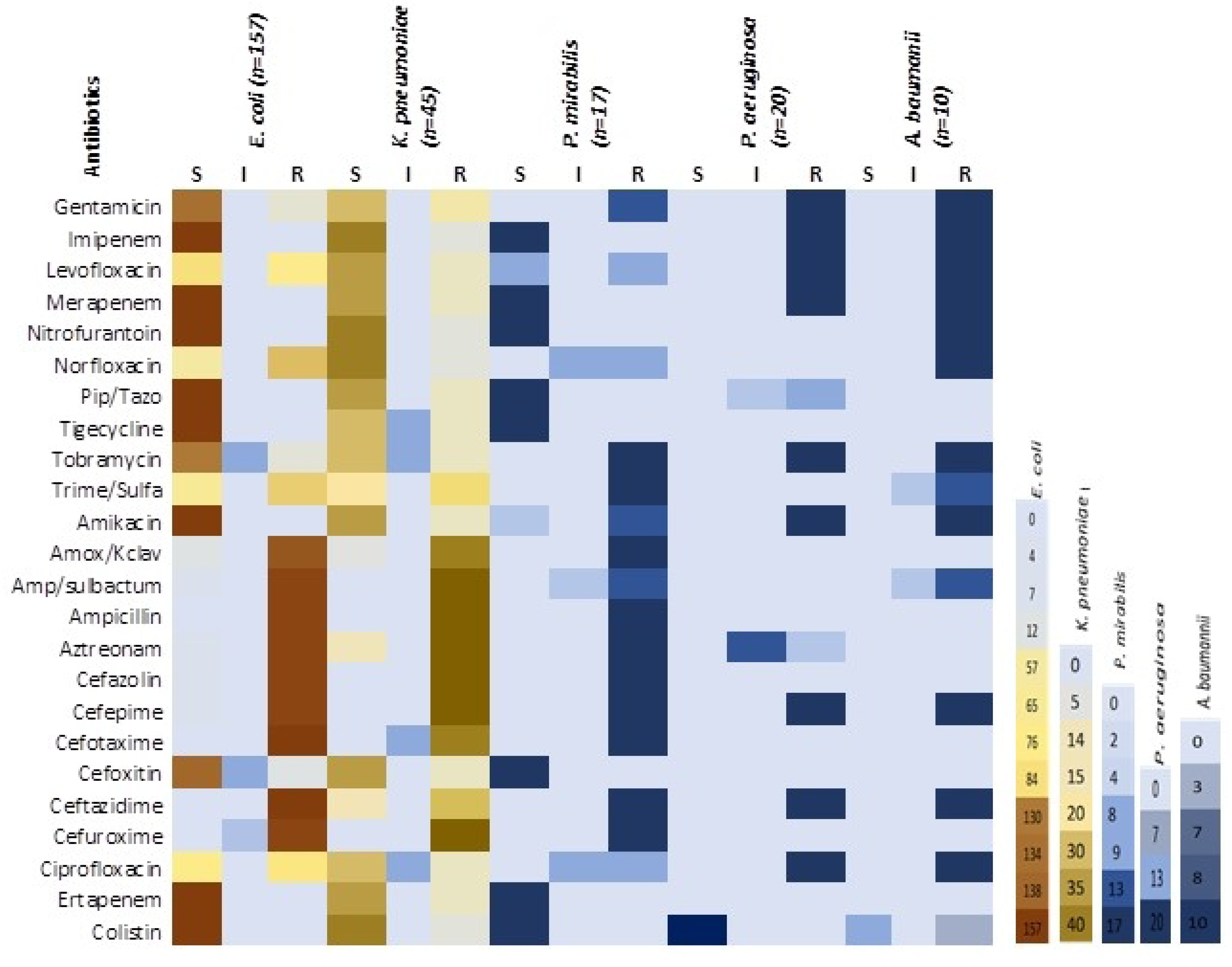
Heatmap of antibiotic sensitivity pattern of *E. coli, K. pneumoniae, P. mirabilis, P. aeruginosa,* and *A. baumannii*.

### Detection of Class A ESBL and Class B & D carbapenemase encoding genes

All the ESBL and carbapenem resistant bacteria (*n*=249) had positive to all the 3 ESBL genes and carbapenem resistant genes *bla_NDM_* & *bla_IMP_* either harbored single or multiple. Among the 249 isolates, the *E. coli* (*n*=157; 63.1%) harbored all the ESBL genes and among them, the *bla_CTX-M_* (*n*=72; 45.9%) was predominantly identified, then *bla_TEM_* (*n*=59; 37.6%) and *bla_SHV_* (*n*=31; 19.7%) respectively. Of 157 *E. coli*, 20.33% (*n*=32) of them co-harbored more than one gene like *bla_CTX-M_*_+*TEM*_ or *bla_CTX-M_*_+*SHV*_ or altogether. In that gene combination, the *bla_CTX-M_*_+*TEM*_ (*n*=15; 9.55%) & *bla_CTX-M_*_+*SHV*_ (*n*=15; 9.55%) were detected in 15 *E. coli* strains and all the 3 genes were identified together in 3.8% of the *E. coli*. Similarly, the *bla_CTX-M_* (40%; *n*=18) was the predominant gene even in the MDR *K. pneumoniae* (*n*=45), followed by the *bla_TEM_* (31.1%; n=14), *bla_SHV_* (31.1%; *n*=14), *bla_IMP_* (13.33%; *n*=6) and *bla_NDM_* (2.22%; *n*=1) respectively. The gene combinations such as either the *bla_CTX-M_*+*_TEM_* (*n*=14; 31.1%) or *bla_CTX-M_*+*_SHV_* (*n*=10; 22.2%) or *bla_TEM_*+*_SHV_* (6.6%; *n*=3) or *bla_TEM_*+*_NDM_*(2.22%; *n*=1) were detected in 62.22% of the *K. pneumoniae.* Likewise in the *P. aeruginosa*, the genes *bla_CTX-M_* and *bla_TEM_*were detected in almost all the isolates. Of which, the *bla_IMP_*was detected in 7 (35%) strains and the *bla_SHV_* was detected in 13 (65%) strains. The gene combination such as *bla_CTX-M_*_+*SHV*_ (5%; *n*=1) and *bla_CTX-M_*_+TEM+*SHV* +IMP_ (5%; *n*=1) were detected in 10% of the organism. In the same manner, the gene *bla_CTX-M_* was detected in all the 17 ESBL positive *P. mirabilis* (100%), the *bla_TEM_* was detected in 8 strains (47.1%) and among the 17 *P. mirabilis* strains, 17.6% (n=3) of them had positive to both the genes (*bla_CTX-M_* _+*TEM*_). The above resistant genes holding bacterial strains were predominantly isolated from the urine and pus samples and the details are presented in Table 5; Fig 5.

**Fig 5.**
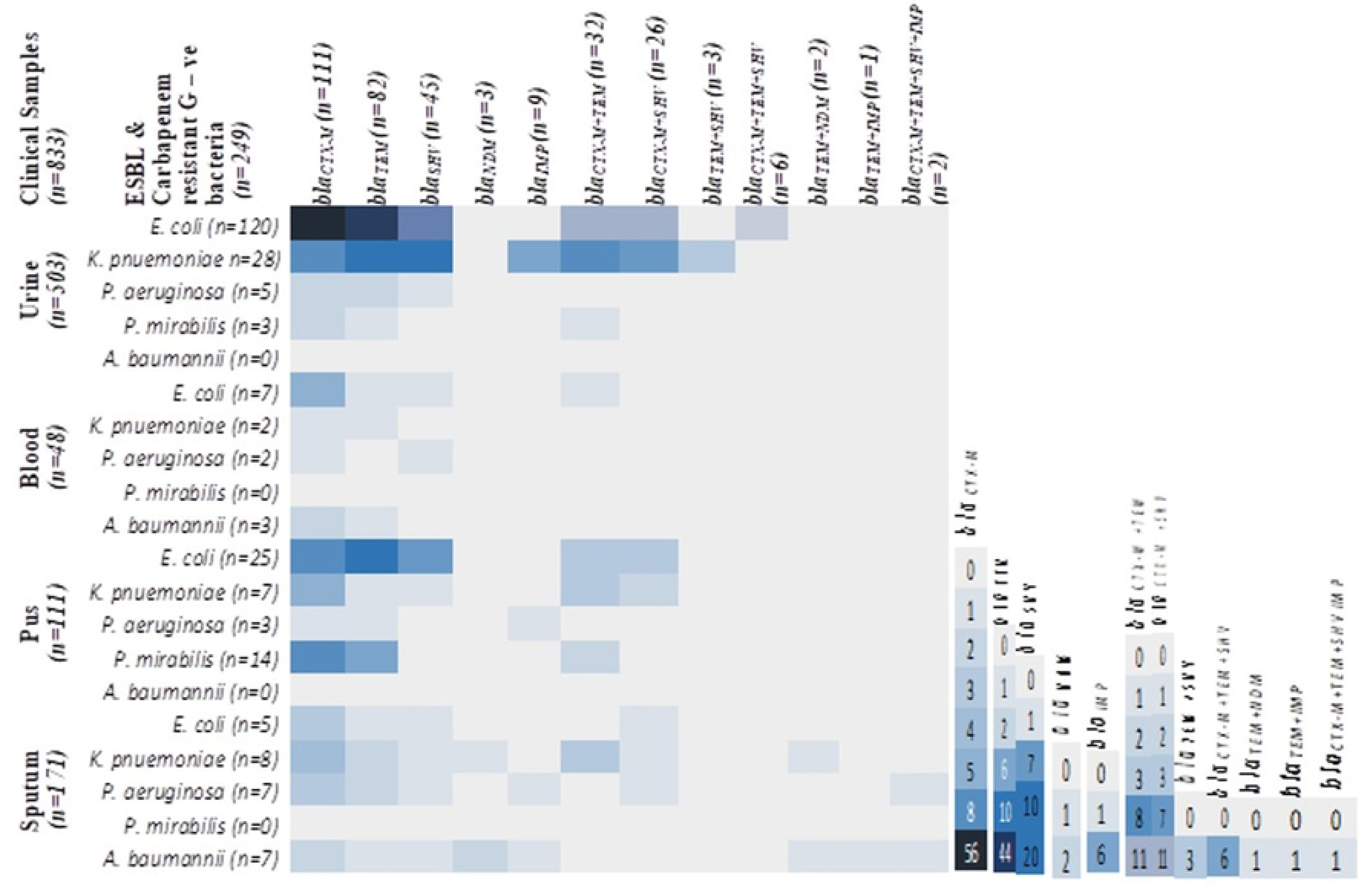
Heatmap of frequency of class A ESBL and class B carbapenemase encoding genes in *E. coli, K. pneumoniae, P. aeruginosa, P. mirabilis* and *A. baumannii* isolated from various clinical samples.

**Table 5.** Number of class A ESBL and class B carbapenemase encoding genes in number of *E. coli, K. pneumoniae, P. aeruginosa, P. mirabilis* and *A. baumannii* isolated from various clinical samples.

| Clinical Samples<br>(n=833) | ESBL & Carbapenem resistant<br>G-ve bacteria<br>n=249<br>(%) |  | Single gene detection |  |  |  |  | Multiple gene detection |  |  |  |  |  |  | Pearson's<br>Chi-square<br>test |
| --- | --- | --- | --- | --- | --- | --- | --- | --- | --- | --- | --- | --- | --- | --- | --- |
|  |  |  | *bla*<sub>CTX-M</sub><br>n=111<br>(%) | *bla*<sub>TEM</sub><br>n=85<br>(%) | *bla*<sub>SHV</sub><br>n=45<br>(%) | *bla*<sub>NDM</sub><br>n=3<br>(%) | *bla*<sub>IMP</sub><br>n=6<br>(%) | *bla*<sub>CTX-M</sub><br>+TEM<br>n=33<br>(%) | *bla*<sub>CTX-M</sub><br>+SHV<br>n=26<br>(%) | *bla*<sub>TEM</sub><br>+SHV<br>n=3<br>(%) | *bla*<sub>CTX-M</sub><br>+TEM<br>+SHV<br>n= 6<br>(%) | *bla*<sub>TEM</sub><br>+NDM<br>n=2<br>(%) | *bla*<sub>TEM</sub><br>+IMP<br>n=1<br>(%) | *bla*<sub>CTX-M</sub><br>+TEM+SHV<br>+IMP<br>n=2<br>(%) |  |
| Urine<br>(n=503) | n=156<br>(31%) | *E. coli*<br>n=120 (76.9%) | 56<br>(46.7%) | 44<br>(36.7%) | 20<br>(16.7%) | 0 | 0 | 11<br>(9.2%) | 11<br>(9.2%) | 0 | 6<br>(5%) | 0 | 0 | 0 | 0.0147 |
|  |  | *K. pneumoniae*<br>n=28 (17.9%) | 8<br>(28.6%) | 10<br>(35.7%) | 7<br>(35.7%) | 0 | 3<br>(21.4%) | 8<br>(28.6%) | 7<br>(25%) | 3<br>(10.7%) | 0 | 0 | 0 | 0 |  |
|  |  | *P. aeruginosa*<br>n=5(3.2%) | 2<br>(40%) | 2<br>(40%) | 1<br>(20%) | 0 | 0 | 0 | 0 | 0 | 0 | 0 | 0 | 0 |  |
|  |  | *P. mirabilis*<br>n=3 (1.9%) | 2<br>(66.7%) | 1<br>(33.3%) | 0 | 0 | 0 | 1<br>(33.3%) | 0 | 0 | 0 | 0 | 0 | 0 |  |
|  |  | *A. baumannii*<br>(n=0) | 0 | 0 | 0 | 0 | 0 | 0 | 0 | 0 | 0 | 0 | 0 | 0 |  |
| Blood<br>(n=48) | n=14<br>(29.2%) | *E. coli*<br>n=7 (50%) | 5<br>(71.4%) | 1<br>(14.3%) | 1<br>(14.3%) | 0 | 0 | 1<br>(14.3%) | 0 | 0 | 0 | 0 | 0 | 0 | 0.7933 |
|  |  | *K. pneumoniae*<br>n=2 (14.3%) | 1<br>(50%) | 1<br>(50%) | 0 | 0 | 0 | 0 | 0 | 0 | 0 | 0 | 0 | 0 |  |
|  |  | *P. aeruginosa*<br>n=2 (14.3%) | 1<br>(50%) | 0 | 1<br>(50%) | 0 | 0 | 0 | 0 | 0 | 0 | 0 | 0 | 0 |  |
|  |  | *P. mirabilis*<br>(n=0) | 0 | 0 | 0 | 0 | 0 | 0 | 0 | 0 | 0 | 0 | 0 | 0 |  |
|  |  | *A. baumannii*<br>n=3 (1.9%) | 2<br>(66.7%) | 1<br>(33.3%) | 0 | 0 | 0 | 0 | 0 | 0 | 0 | 0 | 0 | 0 |  |
| Pus<br>(n=111) | n=49<br>(44.1%) | *E. coli*<br>n=25 (51%) | 8<br>(32%) | 10<br>(40%) | 7<br>(28%) | 0 | 0 | 3<br>(12%) | 3<br>(12%) | 0 | 0 | 0 | 0 | 0 | 0.0054 |
|  |  | *K. pneumoniae*<br>n=7 (14.3%) | 5<br>(71.4%) | 1<br>(14.3%) | 1<br>(14.3%) | 0 | 0 | 3<br>(42.9%) | 2<br>(28.6%) | 0 | 0 | 0 | 0 | 0 |  |
|  |  | *P. aeruginosa*<br>n=3 (6.1%) | 1<br>(33.3%) | 1<br>(33.3%) | 0 | 0 | 1<br>(33.3%) | 0 | 0 | 0 | 0 | 0 | 0 | 0 |  |
|  |  | *P. mirabilis* n=14<br>(28.6%) | 8<br>(57.1%) | 4<br>(42.9%) | 0 | 0 | 0 | 2<br>(14.3%) | 0 | 0 | 0 | 0 | 0 | 0 |  |
|  |  | *A. baumannii*<br>(n=0) | 0 | 0 | 0 | 0 | 0 | 0 | 0 | 0 | 0 | 0 | 0 | 0 |  |
| **Sputum**<br>**(n=171)** | n=30<br>(17.5%) | *E. coli*<br>n=5 (16.66%) | 3<br>(60%) | 1<br>(20%) | 1<br>(20%) | 0 | 0 | 0 | 1<br>(20%) | 0 | 0 | 0 | 0 | 0 | 0.7354 |
|  |  | *K. pneumoniae*<br>n=8 (26.66%) | 4<br>(50%) | 2<br>(25%) | 1<br>(12.5%) | 1<br>(12.5%) | 0 | 3<br>(37.5%) | 1<br>(12.5%) | 0 | 0 | 1<br>(12.5%) | 0 | 0 |  |
|  |  | *P. aeruginosa*<br>n=10 (33.33%) | 3<br>(42.9%) | 5<br>(28.6%) | 1<br>(14.3%) | 0 | 1<br>(14.3%) | 1<br>(14.3%) | 1<br>(14.3%) | 0 | 0 | 0 | 0 | 1<br>(14.3%) |  |
|  |  | *P. mirabilis*<br>(n=0) | 0 | 0 | 0 | 0 | 0 | 0 | 0 | 0 | 0 | 0 | 0 | 0 |  |
|  |  | *A. baumannii*<br>n=7 (23.33%) | 2<br>(28.6%) | 1<br>(14.3%) | 1<br>(14.3%) | 2<br>(28.6%) | 1<br>(14.3%) | 0 | 0 | 0 | 0 | 1<br>(14.3%) | 1<br>(14.3%) | 1<br>(14.3%) |  |

All the *A. baumannii* (*n*=10; 100%) had positive to the gene *bla_CTX-M_* and *bla_TEM_*, 20% (*n*=2) of the strains individually had the gene *bla_NDM_* and *bla_SHV_*, and the *bla_IMP_* was detected in 10% of the *A. baumannii* isolates of the sputum samples. The gene combination *bla_TEM_*_+*NDM,*_ *bla_TEM_*_+*IMP*_ and *bla_CTX-M+TEM+SHV+IMP_* were detected separately in 10% (*n*=1) of the strains. In none of the ESBL and carbapenem producers, the class D carbapenemase gene *bla_OXA-48_* was not detected, and the above-mentioned details are given in Table 6; Fig 6.

**Fig 6.**
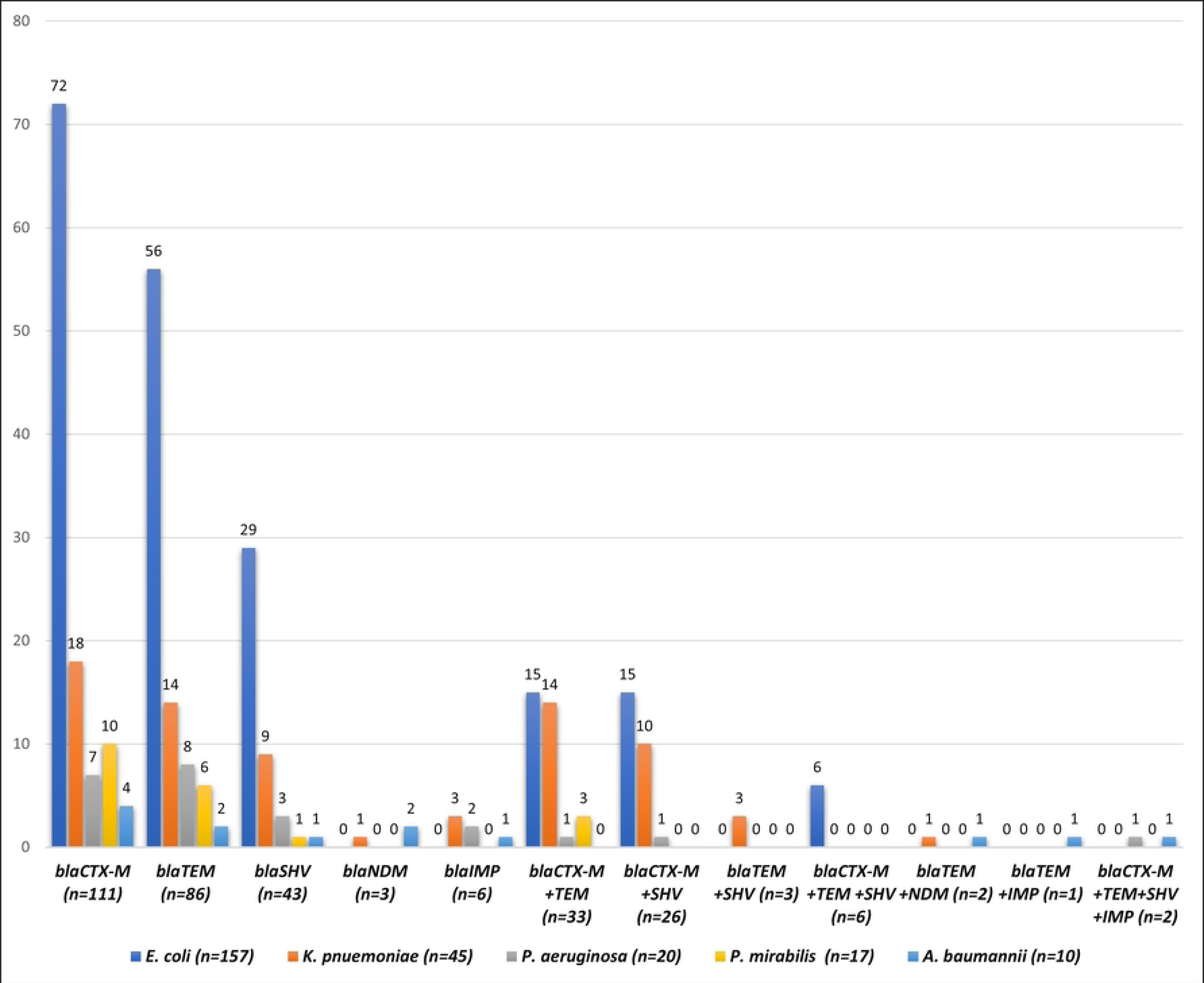
Frequency of detection of class A ESBL and class B & D carbapenemase encoding genes in *E. coli, K. pneumoniae, P. aeruginosa, P. mirabilis* and *A. baumannii*.

**Table 6.**
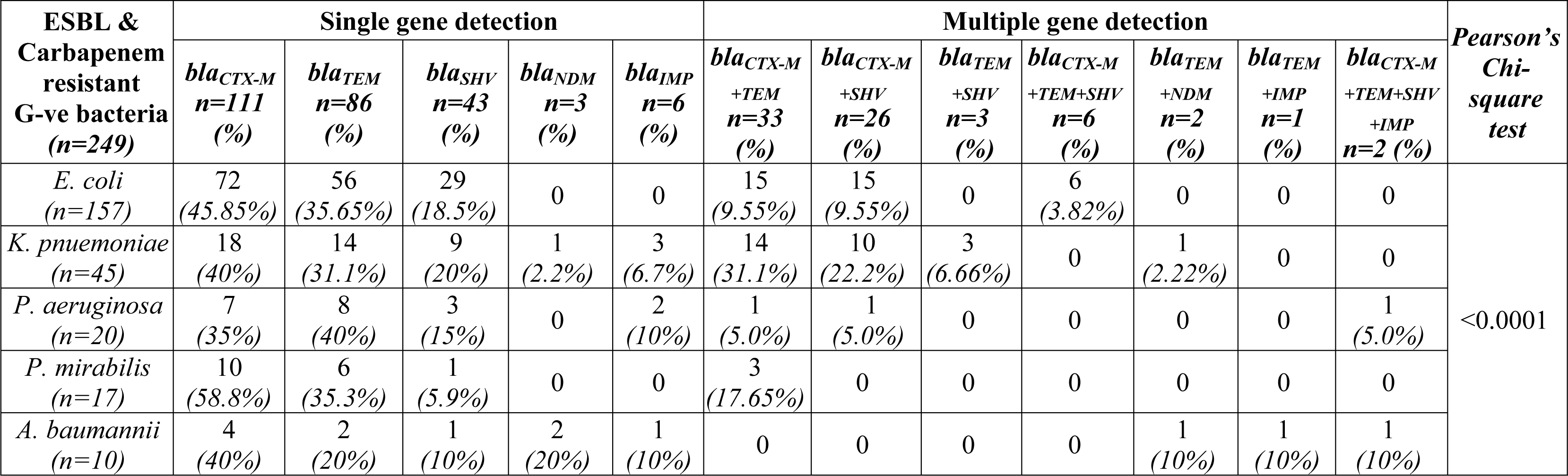
Frequency of detection of class A ESBL and class B carbapenemase encoding genes in *E. coli, K. pneumoniae, P. aeruginosa, P. mirabilis* and *A. baumannii*.

### Genomic sequencing and phylogenetic tree relationships of the *bla_CTX-M_*

The gene *bla_CTX-M_* was isolated and polymerized from the bacteria *E. coli* isolated from the urine sample. Partial gene sequencing of the *bla_CTX-M_* was deposited in the GenBank, NCBI and obtained accession number 2780010. The cladogram of the phylogenetic tree was constructed using 99.74% similarity identification with various *E. coli* & *K. pneumoniae bla_CTX-M_* genes which are already deposited in the GenBank, and the cladogram of the tree is given in Fig 7.

**Fig 7.**
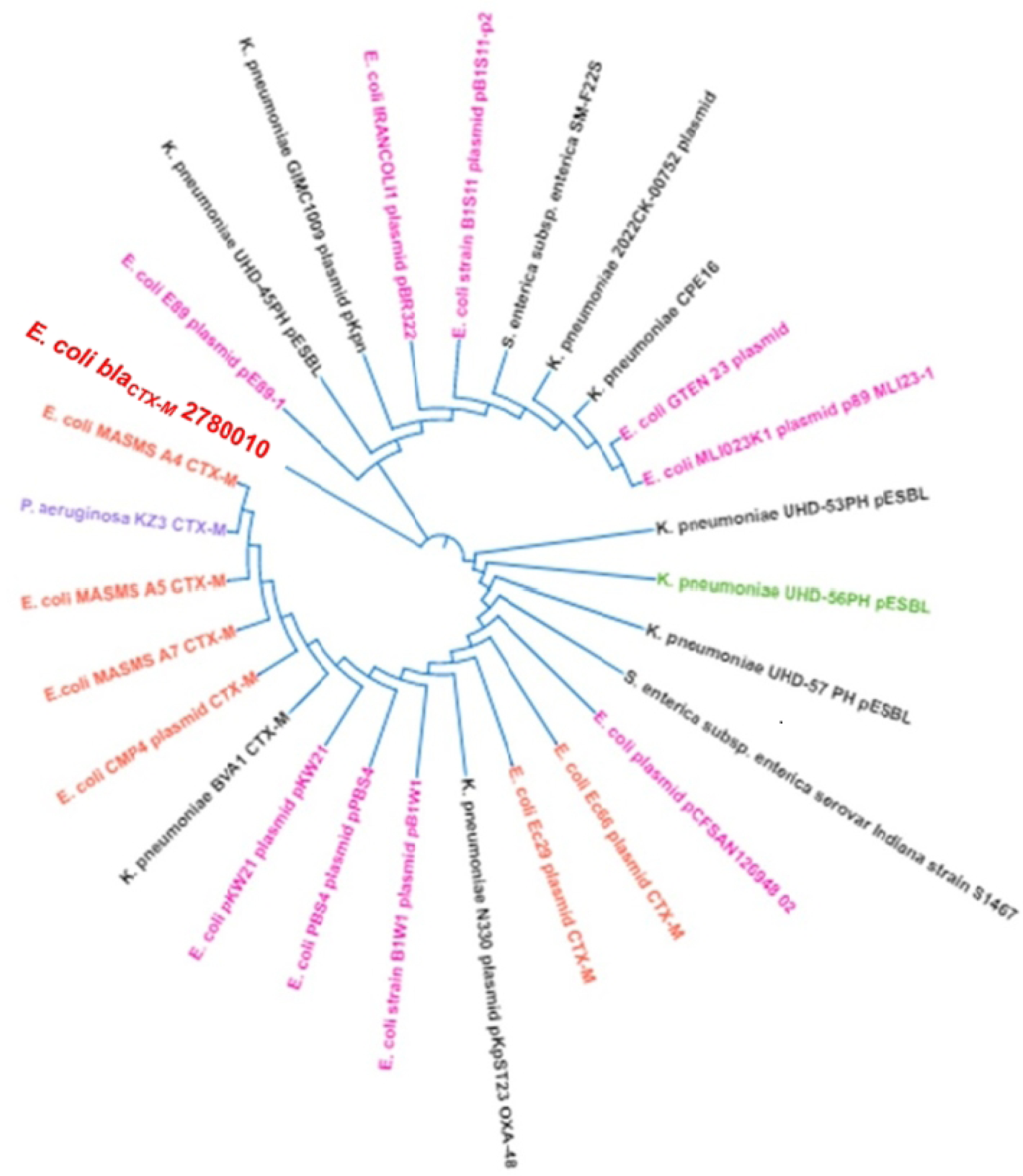
Cladogram of the phylogenetic tree of the class A ESBL gene *bla_CTX-M_* isolated from the ESBL *E. coli*.

## DISCUSSION

Over the recent decades, the spread of ESBL producing *Enterobacteriaceae* has risen as a global concern because of substantial increases in mortality and morbidity and making it difficult to treat infections.[30] Reduced treatment options, complex infections, and costly treatments are some of the major concerns for people infected with ESBL-producing organisms [31]. A major driver of ESBL producing Gram-negative bacteria is the horizontal transfer of mobile genetic elements carrying genes for ESBL and/or carbapenemase enzyme production [32].

The present study collected 3670 various clinical samples, of which the urine samples were predominantly (51.2%) received and had shown the highest bacterial growth rate (49.1%). The bacterial growth rate was significant among the collected samples and the overall positivity rate of the bacterial growth was 29.9%. Similarly, a study was carried out in Abu Dhabi [33] reported that the urine sample was the most common sample type (24.2%), followed by sputum, wound and blood respectively and Mahamat *et al.* stated that in their study the predominant clinical samples were urine (72.5%) than other clinical samples and the highest bacterial growth (70.3%) rate was observed from the urine samples [33, 34]

In this study, 64.3% of the bacterial isolates were gram-negative bacteria and remaining 35.7% were gram positive bacteria (21.5%) and other gram variable bacteria & *Candida* species (14.2%). Among the gram-negative bacteria, the *E. coli* (42.3%) was the most predominant isolates followed by *K. pneumoniae* (19.4%), *P. aeruginosa* (13.5%), *A. baumanii* (7.3%) and *P. mirabilis* (3.3%). Likewise, National AMR Surveillance system of the UAE reported (in 2020) that the *E. coli* was the most commonly (80%) isolated gram-negative bacteria, then *K. pneumoniae* (55%-60%), *P. aeruginosa* (20%), *A. baumannii* (<10%) and *P. mirabilis* (<5%) [35]. In our study, the ESBL *E. coli* had shown 100% resistance to cefotaxime and ceftazidime, similarly, the study was conducted by Al-Agamy *et al.* and Kabrah found 100% resistance to the same drugs by the ESBL *E. coli* [36, 37]. High - medium resistance rates (97.5% - 46.5%) were detected to 11 different antibiotics. The authors Kasim OA (2019), Shaaban *et al.* (2021) & Mohammed *et al* (2022) from Bahrain, Alfaresi *et al.* (2018) & Karlowsky *et al.* (2022) from UAE were found that high-medium resistance to the same drugs in their studies [38–42]. Lower resistance rate to gentamycin (14.7%), tobramycin (12.7%), least resistance to cefoxitin (7.6%) and 100% sensitivity to 8 different antibiotics which are mentioned in Table **4**. Likely the Kasim OA, Perez-Lopez *et al* from Qatar and Al-Tawfiq *et al.* from Saudi Arabia were noted the same results in their studies [38, 43, 44].

The ESBL *K. pneumoniae* were 100% resistant to ampicillin, aztreonam, cefazolin, cefepime and cefuroxime. Multiples studies from different GCC region including UAE reported the pattern of sensitivity to the same drugs [40, 42, 45–47]. Higher resistance rate (89%) was identified on cefotaxime and amoxicillin+clavulanate and this was like the study from Bahrain [38], but lower – medium resistance rate was observed to the above drugs in various studies conducted in Kuwait, UAE, Qatar and Saudi Arabia [37, 43, 47, 48]. The bacteria had shown lower resistance rate (22.2%) to tigecycline, meropenem and other antibiotics and the studies from KSA [49, 50] reported that the resistance rate has increased from 8% to 23.4% almost like the current study. The resistant rate was 11.1% to colistin & imipenem and the rate has increased from 0% to 11.1% during the past 5 years in UAE [41]. The *P. mirabilis* was 100% resistance and 100% sensitive to 10 and 8 different antibiotics respectively and are mentioned in the Table 4 and high – medium resistance rate (76.5% - 53%) had identified to gentamycin, amikacin, ampicillin/sulbactam, ciprofloxacin and norfloxacin. Rather JI *et al*. stated the same finding on the *P. mirabilis* [51]. But in contrast, the current resistant rate was slightly higher than the study reported from China [52].

The bacteria *P. aeruginosa* were 100% resistant to 9 different drugs and 100% susceptible to colistin and medium – low resistance rate was noted to piperacillin/tazobactam (65%) and aztreonam (35%). In contrast to our study, the studies from Sudan [53] and Egypt [54] observed that the bacteria had low resistance rate and high susceptible to the above-mentioned drugs. In the case of *A. baumannii*, the bacteria were 100% resistant to 11 drugs which are given in the table 4, higher resistance rate (70% - 80%) was observed on the ampicillin/sulbactam & trimethoprim/sulfamethoxazole and 30% resistance to colistin. This was very high resistance compared with studies from Oman [55] and Iran [56]. Resistance to the colistin is rapidly emerging among the MDR *A. baumannii* and *K. pneumoniae,* which has become a global burden.

At present, more than 300 different ESBL variants have been identified, the CTX-M, SHV and TEM are the most reported in several areas of the world [57]. The UAE is considered among the GCC countries with the moderate rate of ESBL production among the *Enterobacteriaceae* [41, 58].

In the present study, all the 3 ESBL genes were detected in all the bacterial isolates. Among them, the *bla_CTX-M_* was predominantly harbored, mostly in the *E. coli* (45.9%), *P. mirabilis* (58.8%), *K. pneumoniae* (42.2%), *A. baumannii* (40%) and *P. aeruginosa* (35%). Previous studies from different countries in the GCC region, stated that the *bla_CTX-M_* was the dominant gene among ESBL *Enterobacterales* (100%), *E. coli* (55.5%-100%) and *K. pneumoniae* (32.1% - 100%) across different sample types and clinical settings [41,45,48,59,60]. After that, *bla_TEM_* and *bla_SHV_* were the next commonly detected genes and mainly in the *E. coli* (35.6% & 18.5%) and *K. pneumoniae* (16.4% & 26.7%). Similarly in the UAE, Alfaresi *et al.* stated that the *bla_CTX-M_* was the dominant gene followed by *bla_TEM_* and *bla_SHV_*in their study and Eltai *et al.* found the same dominant gene similarity in their study [41, 59]. In contrast to our results, a study from Iraq stated that the *bla_TEM_* was the most predominant gene in the *Enterobacterales* then followed by *bla_CTX-M_* & *bla*_SHV_.[61] Likewise, Bajpai *et al.* also reported the same as the most prevalent gene was bla*_TEM_* and the *bla_SHV_* [62] and in Zaki study the *bla_SHV_* (61.22%) was the most detected gene [63].

The carbapenemase encoding gene *bla_IMP_* was detected in 13.3% of *K. pneumoniae* isolated from the urine samples and in 10% of *A. baumannii* from sputum & 10% of *P. aeruginosa* from both the samples. Like this, 20% of the *A. baumannii* and 2.22% of *K. pneumoniae* harbored the *bla_NDM_* and they were isolated from the sputum samples. Almost 94% of them were positive to the *bla_IMP_* in Bharain [64] but 6% of the *A. baumannii* were positive to the *bla_NDM_*. 8.7% of the *K. pneumoniae* were positive to the *bla_IMP_* in 2019, Saudi Arabia [46], Perez-Lopez *et al.* [43] reported that 3% of *E. coli* & 2.4% of the *E. coli* were positive to the *bla_NDM_*and *bla_OXA-48_* respectively, and 12.9% & 3.2% of the *K. pneumoniae* were harbored the same genes. Moreover, the gene *bla_OXA-48_*was commonly identified among the *Enterobacteriaceae* in other countries including like Saudi Arabia and Egypt [65–67]. However, in our study, none of the ESBL and the carbapenem resistant bacteria harbored the gene *bla_OXA-48_*. But in contrast to our study, Kamel NA *et al.* from Egypt reported that 6% *E. coli*, 9.4% *Klebsiella* spp, & 3.1% of each *P. aeruginosa* and *A. baumannii* were positive to the *bla_NDM_* and none of the gram-negative bacteria were carried the *bla_IMP_* in their study, but they were positive to the *bla_OXA-48_.*[68]

The highest co-existence patterns were *bla_CTX-M+TEM_* and *bla_CTX-M+SHV_* represented by *E. coli* (9.6% of each), *K. pneumoniae* (31.1% & 22.2%) & *P. aeruginosa* (15% of each). In 17.65% of the *P. mirabilis* the only co-existence gene was *bla_CTX-M+TEM_.* The lowest co-existence patterns were *bla_TEM+SHV_* & *bla_CTX-M+TEM+SHV_*represented by *K. pneumoniae* (6.67%) and *E. coli* (3.82%) respectively. Similarly, the highest co-existence of the gene *bla_CTX-M+TEM_* (21%) was found in the ESBL *E. coli* & *K. pneumoniae* from Qatar [59], Saudi Arabia [45, 69] and followed by the least co-existence of the genes *bla_TEM+SHV,_ bla_CTX-M+SHV_* & *bla_CTX-M+TEM+SHV_* were detected in their same studies, but in contrast, the Moglad *et al*, Badger-Emeka *et al* and Sid Ahmed *et al* found that the highest co-harbored genes were *bla_CTX-M+TEM+SHV_* (50% & 24.7%), *bla_CTX-M+SHV_* (50% & 10.1%) found in the *E. coli*, *K. pneumoniae* and other *Enterobacterales* in their studies. [70,71] Likewise, the lowest co-expression of the genes *bla_TEM+NDM_*, *bla_TEM+IMP_* and *bla_CTX-M+TEM+SHV+IMP_* were also detected in 2.22% *K. pneumoniae,* 5% *P. aeruginosa* and 10% of each *A. baumannii* to each gene combination. In such a way, Elbadawi HS e*t al.* found in their study that the lowest co-expression of the ESBL and carbapenemase producing genes were noticed in the *K. pneumoniae, A. baumannii, E. coli* and *P. aeruginosa*. [72]

## CONCLUSION

The current findings confirm UAE to have a slightly high prevalence of class A ESBL and class B carbapenemase encoding genes. According to the study the gene *bla_CTX-M_*, *bla_TEM_* and *bla_SHV_*were the most detected in the *E. coli* and *K. pneumoniae* especially isolated from the urine samples. The highest percentage of the bacteria *K. pneumoniae* harbored all the class A ESBL & class B carbapenemase encoding genes and, the multiple two gene combinations like *bla_CTX-M+TEM_*, *bla_CTX-M+SHV_*, *bla_TEM+SHV_* & *bla_TEM+NDM_* as well and then *A. baumannii* also harbored all the class A ESBL and class B carbapenemase encoding genes and also they co-harbored the genes *bla_TEM+NDM_*, *bla_TEM+IMP_*, *bla_CTX-M+TEM+SHV+IMP._* The current results emphasize the significance of rational antibiotic therapy and ongoing stringent surveillance and infection control strategies to successfully curb the spread of these ESBL and carbapenemase encoding genes in the gram – ve bacteria and become multidrug resistant pathogens.

### Limitations of the study

As of all other studies this study also reported few limitations as follow.

a. The study samples received from various hospitals, clinics and Thumbay hospitals located in and around the northern emirates of UAE (Fujairah, Umm-Al-Quwain, Ajman, and Sharjah) and not received the samples from other emirates (Ras-Al-Khaimah, Dubai & Abu Dhabi).
b. The class D carbapenemase encoding gene *bla_OXA-48_* was detected in none of the ESBL and carbapenem resistant gram – ve bacterial pathogens even in *A. baumannii*. In this study, partial sanger sequencing of the predominant *gene blaCTX-M* was done but the other next predominant class A genes *bla_TEM_* & *bla_SHV_* could not be able to do the sanger sequencing.

## Supporting Information

**Supplementary table 1a** - Test wells on the MicroScan Gram-negative panels (Neg Breakpoint Combo 50).

**Supplementary table 1b** - Antibiotic sensitivity test wells on the MicroScan Gram-negative panels (Neg Breakpoint Combo 50).

**S1 File.** For each variable of interest, give sources of data and details of methods of assessment (measurement). Describe comparability of assessment methods if there is more than one group.

**S2 File.** (a) Report numbers of individuals at each stage of study—e.g., numbers potentially eligible, examined for eligibility, confirmed eligible, included in the study, completing follow-up, and analyzed.

**S3 file.** Flow diagram of the study.

**S4 File.** Cross-sectional study—Report numbers of outcome events or summary measures.

**S5 File.** Cautious overall interpretation of results considering objectives, limitations, multiplicity of analyses, results from similar studies, and other relevant evidence.

## Data Availability

Not applicable

## Acknowledgments

We would like to acknowledge the efforts of Physicians, Microbiologists and other relevant staffs of Thumbay Laboratory and Thumbay University Hospital, Ajman, U.A.E for collecting the gram-negative bacterial pathogens and data and conducting this study. Also want to express our gratitude to the College of Medicine, GMU for the constant support to conduct this research.

## Author contributions

**Conceptualization:** Nazeerullah Rahamathullah

**Data curation:** Nazeerullah Rahamathullah, Premalatha Ragupathi, Vaneezeh Khamisani, Aisha Fadila Sadiq, Mariam Aylu Mobiddo.

**Formal analysis:** Premalatha Ragupathi, Vaneezeh Khamisani, Aisha Fadila Sadiq, Mariam Aylu Mobiddo.

**Investigation:** Nazeerullah Rahamathullah, Premalatha Ragupathi, Vaneezeh Khamisani, Aisha Fadila Sadiq, Mariam Aylu Mobiddo.

**Methodology:** Nazeerullah Rahamathullah, Premalatha Ragupathi, Vaneezeh Khamisani, Aisha Fadila Sadiq, Mariam Aylu Mobiddo, Alaa Kamian.

**Project administration:** Nazeerullah Rahamathullah, Nasir Parwaiz.

**Software:** Nazeerullah Rahamathullah, Premalatha Ragupathi, Vaneezeh Kasmian.

**Supervision:** Nazeerullah Rahamathullah, Mohammad Shahid Akhtar, Nasir Parwaiz.

**Validation:** Nazeerullah Rahamathullah, Sovan Bagchi.

**Writing – original draft:** Premalatha Ragupathi, Vaneezeh Khamisani, Aisha Fadila Sadiq, Mariam Aylu Mobiddo, Nazeerullah Rahamathullah, Sovan Bagchi.

**Writing – review & editing:** Nazeerullah Rahamathullah, Sovan Bagchi.

